# Precision phenotyping from routine laboratory parameters for machine learning out-of-hospital survival prediction using 4D time-dependent SHAP plots in an all-comers prospective PCI registry

**DOI:** 10.1101/2024.08.31.24312888

**Authors:** Paul-Adrian Călburean, Anda-Cristina Scurtu, Paul Grebenisan, Ioana-Andreea Nistor, Victor Vacariu, Reka-Katalin Drincal, Ioana Paula Sulea, Tiberiu Oltean, László Hadadi

**Affiliations:** Emergency Institute for Cardiovascular Diseases and Transplantation Târgu Mureş, Târgu Mureş, Romania; University of Medicine, Pharmacy, Science and Technology “George Emil Palade” of Târgu Mureş, Târgu Mureş, Romania

**Keywords:** Machine learning, coronary artery disease, percutaneous coronary intervention, survival analysis

## Abstract

**Introduction:** Out-of-hospital mortality in coronary artery disease (CAD) is particularly high and established adverse event prediction tools are yet to be available. Our study aimed to investigate whether precision phenotyping can be performed using routine laboratory parameters for the prediction of out-of-hospital survival in a CAD population treated by percutaneous coronary intervention (PCI).

**Materials and methods:** All patients treated by PCI and discharged alive in a tertiary center between January 2016 – December 2022 that have been included prospectively in the local registry were analyzed. 115 parameters from the PCI registry and 266 parameters derived from routine laboratory testing were used. An extreme gradient-boosted decision tree machine learning (ML) algorithm was trained and used to predict all-cause and cardiovascular-cause survival.

**Results:** A total of 7186 PCI hospitalizations for 5797 patients were included with more than 610.000 laboratory values. All-cause and cardiovascular cause mortality was 17.5% and 12.2%, respectively, during a median follow-up time of 1454 (687 – 2072) days. The integrated area under the receiver operator characteristic curve for prediction of all-cause and cardiovascular cause mortality by the ML on the validation dataset was 0.844 and 0.837, respectively (all p<0.001). The integrated area under the precision-recall curve for prediction of all-cause and cardiovascular cause mortality by the ML on the validation dataset was 0.647 and 0.589, respectively (all p<0.001).

**Conclusion:** Precise survival prediction in CAD can be achieved using routine laboratory parameters. ML outperformed clinical risk scores in predicting out-of-hospital mortality in a prospective all-comers PCI population.

## Introduction

Extensive research has been dedicated to the prediction of adverse events in the coronary artery disease (CAD) population. Numerous clinical, angiographic, and combined risk scores were developed to estimate survival, such as Global Registry of Acute Coronary Events (GRACE) score, Age, Creatinine, and Ejection fraction (ACEF) score, thrombolysis in myocardial infarction (TIMI) score and Synergy Between PCI with TAXUS and Cardiac Surgery (SYNTAX) score.^1–4^ However, in many available risk scores the prediction performance is mainly driven by high accuracy in predicting in-hospital death, since the presence of very high-risk clinical features almost invariably leads to in-hospital death (e.g., cardiogenic shock, acute pulmonary edema, resuscitated cardiac arrest, mechanical complications of acute myocardial infarction, severe anemia or acute kidney injury), while out-of-hospital death is more difficult to predict. Moreover, long-term survival in CAD is less determined by the complexity of coronary anatomy.^5^ Thus, more research is needed to find robust predictors of impaired survival in the CAD population, especially after hospital discharge.

Phenotype, initially defined as “the observable traits of an organism”, nowadays includes clinical and paraclinical characteristics of a patient (e.g., diagnostic imaging, laboratory parameters, electrocardiographic changes).^6^ According to the patient’s phenotypic abnormalities, both diagnostic and prognostic assessments can be made.^6^ Precision phenotyping is a concept that implies a comprehensive analysis of a wide panel of clinical parameters usually using artificial intelligence (AI) techniques which can quantify even slight changes in a patient’s phenotype and offer a precise prognostic estimate. Precision phenotyping has been previously proposed using clinical, ultrasound, computed tomography, or big “-omic” data.^7–9^ Whether a precise survival estimate can be given using a spectrum of laboratory parameters routinely measured in clinical practice is yet to be proven.

The aim of the present study was (1) to train an ML model to predict out-of-hospital all-cause and cardiovascular cause mortality using a wide panel of laboratory parameters, (2) to identify predictors of out-of-hospital adverse events, and (3) to compare the predictive performance of ML model with clinical risk scores relevant for CAD population, in an all-comers patient population treated by PCI in a Romanian tertiary cardiovascular center.

## Materials and methods

### Study population

The study design is illustrated in Figure 1. All patients treated by PCI in the Emergency Institute for Cardiovascular Diseases and Transplantation of Târgu Mureş have been prospectively included at discharge in the local PCI Registry of the Institute since January 2016. For the current analysis, all patients treated between January 2016 – December 2022 were included, in order to have a minimum of 6 months of follow-up. Subjects were analyzed by PCI hospitalization (hospitalization in which PCI was performed). Exclusion criteria consisted of (1) age less than 18 years old, (2) presence of in-hospital death, or (2) lack of available survival data (e.g., foreign patients). The Registry is accessible online at the website http://pci.cardio.ro/ and is based on the criteria of Cardiology Audit and Registration Data Standards (CARDS) developed by the Department of Health and Children, European Society of Cardiology, Irish Cardiac Society, and the European Commission.^10^ Briefly, the CARDS recommendations address data regarding demographics, relevant medical history, and comorbid conditions, clinical status at hospital admission, PCI indication, affected and treated coronary artery segments, usage of invasive diagnostic or therapeutic devices, procedural complications, medical treatment during hospitalization and at discharge and in-hospital evolution. All the information available regarding all the variables proposed in that document was collected for each included patient, at every PCI. A number of 115 parameters were available from the PCI registry. Moreover, to complete and update the original CARDS recommendations regarding PCI procedures, an additional 266 laboratory parameters were also collected (Supplemental Table 1). Noteworthy, all laboratory data during the PCI hospitalization was acquired from the electronic health record (EHR). Since the timing and number of determinations of laboratory parameters differ between patients and hospitalizations, each laboratory parameter was considered separately first value in hospital, last value before discharge, average value during hospitalization, absolute maximum and minimum value, and number of each separate determination of that certain laboratory parameter (e.g., first creatinine, last creatinine, average creatinine, maximum and minimum creatinine, number of creatinine determinations, Supplemental Table 1). A total of 381 parameters were used for the final analysis. Furthermore, clinical, and angiographic risk scores such as ACEF, GRACE, and SYNTAX score were also calculated for comparison with the ML model’s survival prediction performance.

**Figure 1:**
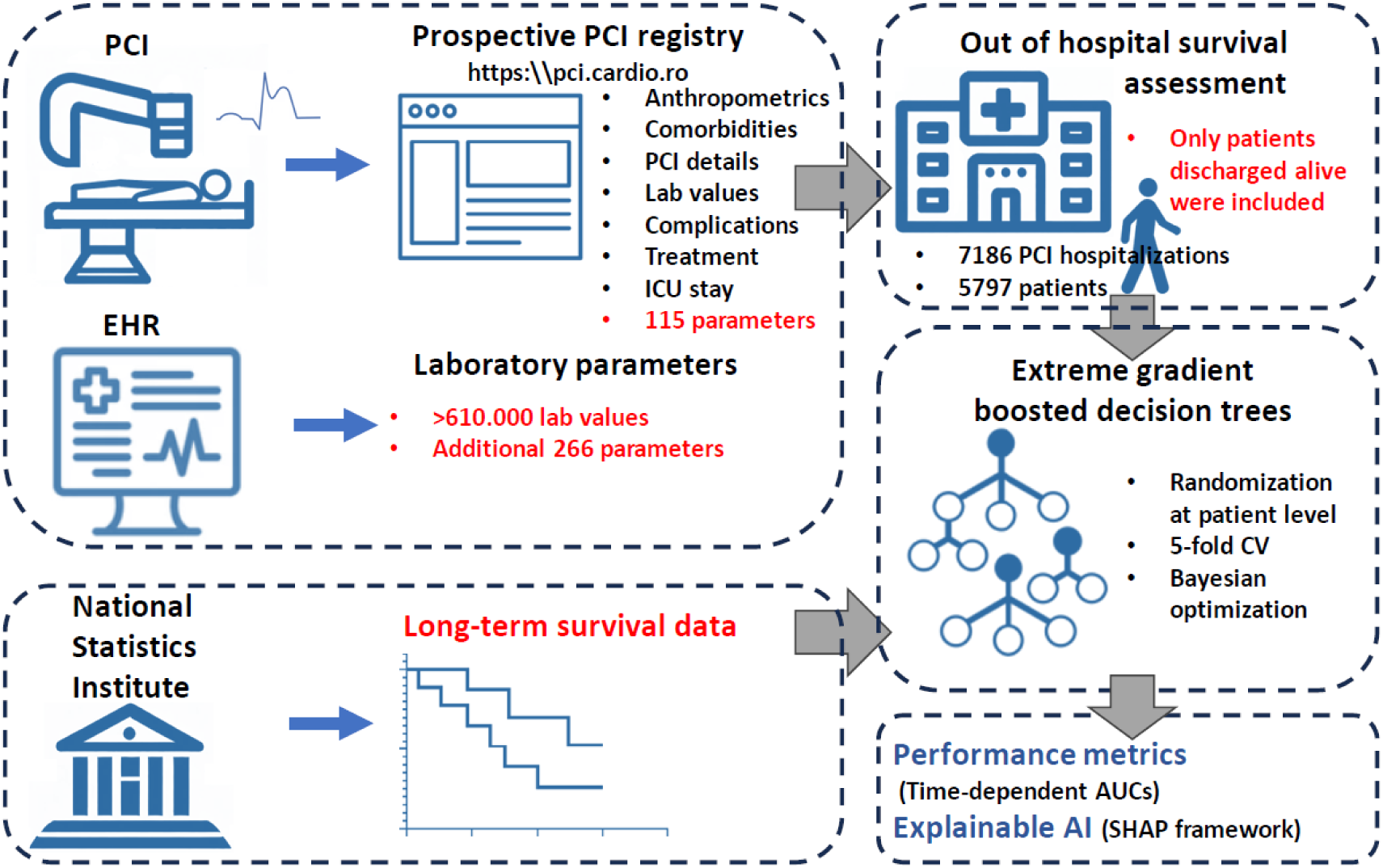
Illustration of the study design.

All patients or their legal representatives (e.g., for critical patients) provided written signed informed consent regarding the PCI procedure and their participation in the study. The study was approved by the ethical committee of our institution (decision number 8646 from 22 December 2015 approved by the Ethical Committee of the Emergency Institute for Cardiovascular Diseases and Transplantation of Târgu Mureş). The protocol was carried out in accordance with the ethical principles for medical research involving human subjects established by the Declaration of Helsinki, protecting the confidentiality of personal information of the patients.

### Follow-up and clinical outcomes

The clinical endpoint of this study was the incidence of out-of-hospital cardiovascular and all-cause mortality. Out-of-hospital mortality was defined as death that occurred after discharge, but not occurring during the initial hospital admission when the PCI procedure was performed. The Romanian National Health Insurance System database supplied mortality rates as of July 2023 for all the patients. For patients who had died during follow-up, the Regional Statistics Office of the Romanian National Institute of Statistics supplied the exact date and cause of death according to the tenth revision of the International Classification of Diseases (ICD-10). If the cause of death belonged to diseases of the circulatory system, then death was considered to be of cardiovascular cause.

### Machine learning

All patients had a minimum of 6 months survival status available and up to 6 years of follow-up. Patients were dichotomized every 6 months into alive/deceased groups and censored events were removed. Afterward, for each timeframe, the dataset was randomly divided into 70% training and 30% validation datasets. To prevent data leaks since a patient could have multiple hospitalizations, each patient was assigned to either a training or validation set. On the 70% training dataset, a 5-fold cross-validation was performed and to prevent data leak between folds, each patient was only assigned to one-fold. An extreme gradient boosted decision tree (XGBoost) algorithm^11^ was evaluated as a binary classifier for predicting both all-cause- and cardiovascular-cause mortality occurred out of hospital, for each timeframe. XGBoost algorithm was implemented in Python version 3.9.13. Hyperparameter optimization was performed by using Bayesian search on 6 years survival status, being the least imbalanced dataset, and those best training parameters were used for all timeframes. The accessibility of underlying supporting scripts is detailed in the data availability section. In order to explain the ML decision process, the open-source Shapley additive explanations (SHAP) framework was used.^12^ Most important features were obtained as follows: (1) SHAP values were obtained for each of the database values at each timeframe, for both all-cause and cardiovascular cause events; (2) at each timeframe, absolute SHAP values were added, and parameters’ importance was considered by the highest sum of absolute SHAP value (e.g., the parameter with the highest absolute SHAP values sum at 5-years cardiovascular survival was the most important); (3) overall most important predictors were obtained by ascending classification of averaged positions at all timeframes for all-cause and cardiovascular cause survival, separately (see supporting code). Illustration of time-dependent SHAP values for a certain parameter on a specified survival (e.g., time-dependent SHAP values for age impact on all-cause survival) was performed as follows: (1) SHAP values were obtained for every value of that parameter at each timeframe on the specified survival; (2) for the specified parameter, ten percentile intervals were obtained according to the minimum and maximum value; (3) SHAP values were averaged for each percentile at each timeframe; (4) 3D mesh plot was illustrated using the ten percentiles of that parameter, all investigated timeframes and averaged SHAP values for that percentile; (5) in order to correlate averaged SHAP values with actual observed survival, the now 4D mesh plot was colored by averaged survival for each parameter percentile at each timeframe (see supporting code).

### Statistical analysis

A significance level α of 0.05 and a 95% confidence interval (CI) were considered. Continuous variables were evaluated for normal distribution using the Shapiro-Wilk test. Continuous variables with parametric distributions were reported as mean ± standard deviation and compared using non-paired Student’s t-test, while continuous variables with non-parametric distributions and discrete variables were reported as median (interquartile range) and compared using Mann Whitney test. Categorical variables were reported as absolute and relative frequencies and compared using Fisher’s exact test for clinical parameters with frequencies less than 5 and the Chi^2^ test otherwise. For baseline clinical characteristics, univariate Cox proportional hazard was used to predict the association in the form of hazard ratio (HR) between observed survival and an independent categorical variable. All continuous variables were dichotomized related to the median to facilitate comparability of HR and CI between variables. Statistical analysis was performed using R version 4.1.1 and R Studio version 1.4.17. Performance on survival prediction was measured by integrating the time-dependent area under the curve of receiver-operator characteristic (AUC-ROC), the area under the curve of precision-recall (AUC-PR), Matthews correlation coefficient (MCC), F1 score, and Brier score for a specified survival. While for AUC-ROC the baseline is constant at 0.5, in the case of AUC-PR, the baseline is determined by positive cases per total cases ratio.^13^ Thus, a baseline-corrected AUC-PR was calculated by subtracting the baseline value from the AUC-PR value. MCC and F1 score calculation require dichotomization from probability for ML model and score value for clinical risk scores into predicted deceased or alive, which was performed using Youden’s method for cut-off calculation.

## Results

### Study population

A total of 7186 PCI hospitalizations for 5797 patients were included in the present study, with a total of 611309 laboratory values. Of those patients, 4062 (70.07%) were male sex, the median age was 65.0 (57.2-71.8) years and the median BMI was 28.3 (25.9-31.8) kg/m^2^. Complete clinical characteristics of the studied patients are reported in Table 1 and Supplemental Table 1. Complete clinical characteristics of the studied PCI hospitalizations are reported in Table 2 and Supplemental Table 2. A total of 1017 (17.5%) patients died of all causes during a median follow-up time of 1454 (687 – 2072) days. When censoring events of non-cardiovascular cause, a total of 710 (12.2%) patients died of cardiovascular causes during a median follow-up time of 1544 (748– 2155) days. Cumulative per-patient mortality incidence is illustrated in Figure 2A and cumulative per-PCI mortality incidence is illustrated in Figure 2B. A total of 115 clinical parameters were available from the PCI registry. A median of 114 (110-115) parameters were available for each patient as some parameters were missing or not applicable (e.g., time from symptom onset to PCI, which is measured in acute but not in chronic coronary syndrome). A total of 611309 laboratory values were analyzed for all 7186 hospitalizations. Per hospitalization, a median of 78 (65-116) laboratory values were available. For each laboratory parameter, the first and last in-hospital value, maximum and minimum value during hospitalization, averaged value during hospitalization, and number of determinations for that parameter during hospitalization were calculated, therefore 266 additional parameters were analyzed.

**Figure 2:**
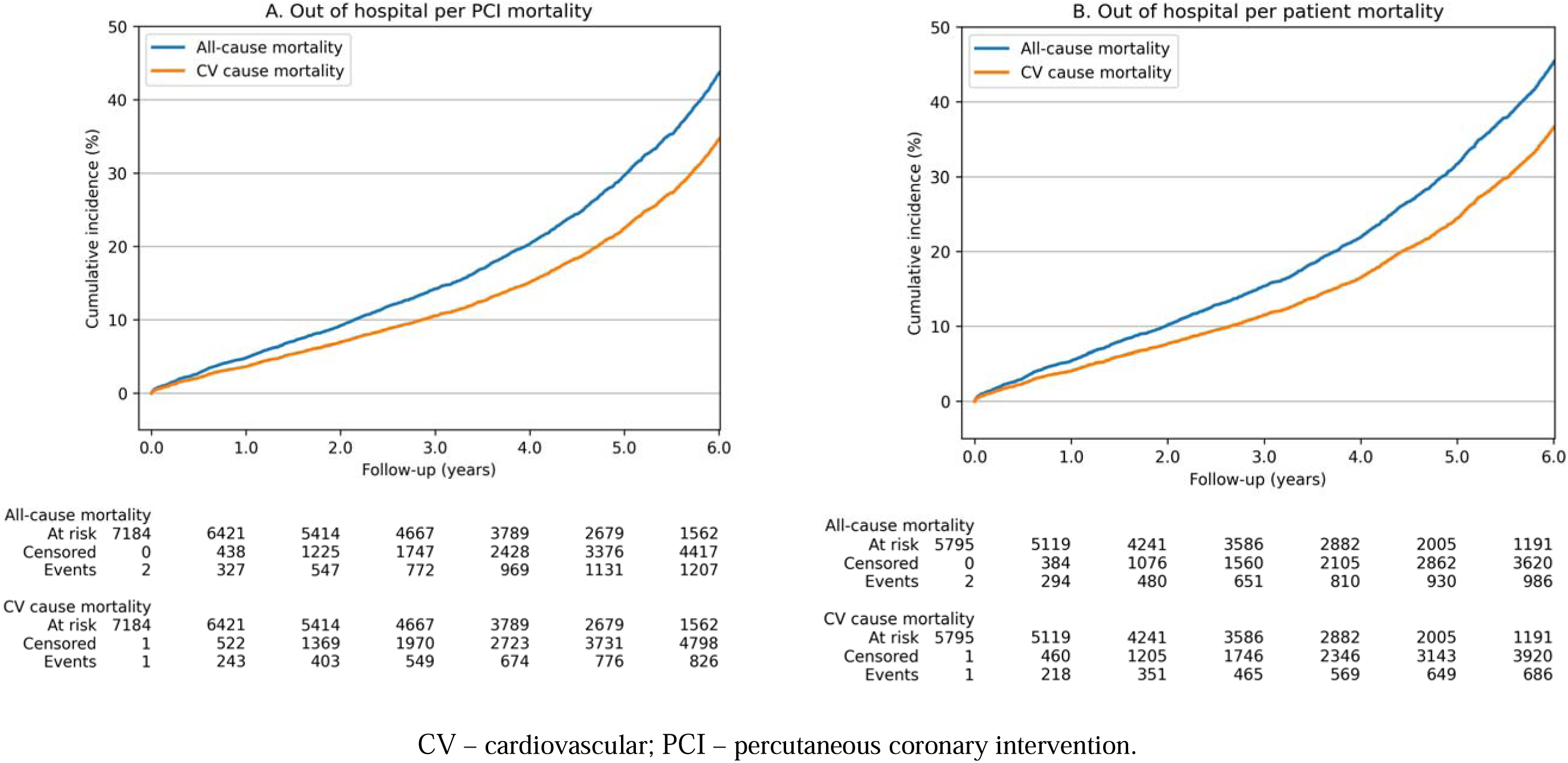
Observed survival in the studied population.

**Table 1:** Baseline characteristics of the studied parameters on a per patient analysis.

| Parameter | All patients<br>(n=5797) | All-cause survival |  |  | CV-cause survival |  |  |
| --- | --- | --- | --- | --- | --- | --- | --- |
|  |  | HR | 95%CI | p | HR | 95%CI | P |
| Male sex | 4062 (70.07%) | 1.04 | 0.91-1.19 | 0.6 | 0.99 | 0.85-1.17 | 0.93 |
| Age (years) | 65 (57-71) | 2.56 | 2.24-2.92 | $<10^{-42}$ | 2.64 | 2.25-3.1 | $<10^{-31}$ |
| BMI (kg/m<sup>2</sup>) | 28 (25-31) | 0.89 | 0.77-1.02 | 0.1 | 0.87 | 0.74-1.04 | 0.13 |
| History of MI | 1140 (19.67%) | 1.16 | 1.01-1.33 | 0.04 | 1.24 | 1.05-1.47 | $<10^{-2}$ |
| CABG | 154 (2.66%) | 1.06 | 0.75-1.5 | 0.73 | 1.13 | 0.76-1.68 | 0.56 |
| Previous PCI | 1284 (22.15%) | 0.89 | 0.77-1.03 | 0.13 | 0.89 | 0.75-1.06 | 0.18 |
| Stroke/TIA | 386 (6.66%) | 2.13 | 1.76-2.57 | $<10^{-14}$ | 2.15 | 1.71-2.69 | $<10^{-10}$ |
| Atrial fibrillation | 695 (11.99%) | 2.14 | 1.84-2.5 | $<10^{-22}$ | 2.33 | 1.95-2.78 | $<10^{-19}$ |
| PAD | 607 (10.47%) | 2.02 | 1.73-2.35 | $<10^{-18}$ | 1.83 | 1.52-2.22 | $<10^{-9}$ |
| CKD | 415 (7.16%) | 3.46 | 2.93-4.09 | $<10^{-47}$ | 3.68 | 3.02-4.47 | $<10^{-38}$ |
| VHD | 332 (5.73%) | 2.16 | 1.78-2.62 | $<10^{-14}$ | 2.35 | 1.88-2.94 | $<10^{-13}$ |
| DCM | 481 (8.30%) | 1.88 | 1.58-2.25 | $<10^{-11}$ | 2.23 | 1.82-2.73 | $<10^{-14}$ |
| COPD | 531 (9.16%) | 2.4 | 2.05-2.82 | $<10^{-26}$ | 2.58 | 2.14-3.11 | $<10^{-22}$ |
| Diabetes Mellitus | 1072 (18.49%) | 1.54 | 1.35-1.77 | $<10^{-9}$ | 1.51 | 1.29-1.78 | $<10^{-6}$ |
| HBP | 2873 (49.56%) | 1.19 | 1.05-1.36 | $<10^{-2}$ | 1.21 | 1.04-1.42 | 0.01 |
| Dyslipidaemia | 2236 (38.57%) | 0.97 | 0.86-1.1 | 0.67 | 0.93 | 0.8-1.08 | 0.32 |
| LVEF (%) | 50 (40-55) | 0.46 | 0.4-0.54 | $<10^{-21}$ | 0.41 | 0.34-0.5 | $<10^{-18}$ |
| NTproBNP(pg/ml) | 934(212-2831) | 2.94 | 1.66-5.21 | $<10^{-3}$ | 1.82 | 0.98-3.36 | 0.06 |
| Creatinine(mg/dL) | 0.91 (0.79-1.1) | 1.81 | 1.59-2.05 | $<10^{-19}$ | 1.97 | 1.69-2.3 | $<10^{-17}$ |
| HGB (g/dL) | 13.8 (12.7-14) | 0.55 | 0.49-0.63 | $<10^{-18}$ | 0.55 | 0.47-0.64 | $<10^{-13}$ |
| PLT ($\times 10^3/\mu\text{L}$) | 227 (188-272) | 0.99 | 0.88-1.12 | 0.89 | 0.95 | 0.82-1.1 | 0.5 |
| WBC ($\times 10^3/\mu\text{L}$) | 8.3(6.7-10.4) | 1.25 | 1.1-1.42 | $<10^{-3}$ | 1.31 | 1.13-1.52 | $<10^{-3}$ |
| STEMI | 1574 (27.15%) | 1.26 | 1.1-1.45 | $<10^{-3}$ | 1.31 | 1.12-1.55 | $<10^{-2}$ |
| NSTE-MI | 517 (8.92%) | 1.64 | 1.35-1.98 | $<10^{-6}$ | 1.73 | 1.39-2.16 | $<10^{-5}$ |
| UA | 1064 (18.35%) | 0.9 | 0.76-1.05 | 0.18 | 0.81 | 0.67-0.99 | 0.04 |
| CCS | 2642 (45.58%) | 0.75 | 0.66-0.85 | $<10^{-5}$ | 0.75 | 0.64-0.87 | $<10^{-3}$ |

BMI – body mass index; CABG – coronary artery bypass graft; CCS – chronic coronary syndrome; CI – confidence interval; CKD – chronic kidney disease; COPD – chronic obstructive pulmonary disease; CV – cardiovascular; DCM – dilated cardiomyopathy; HGB – haemoglobin; HBP – high blood pressure; LVEF – left ventricular ejection fraction; MI – myocardial infarction; PAD – peripheral artery disease; PCI – percutaneous coronary intervention; PLT – platelet count; NSTE-MI – non-ST segment elevation acute MI; STEMI – ST segment elevation acute MI; TIA – transient ischemic attack; UA – unstable angina; VHD – valvular heart disease; WBC – white blood cells count.

**Table 2:** Baseline characteristics of the studied parameters on a per PCI analysis.

| Parameter | All PCIs<br>(n=7186) | All-cause survival |  |  | CV-cause survival |  |  |
| --- | --- | --- | --- | --- | --- | --- | --- |
|  |  | HR | 95%CI | p | HR | 95%CI | p |
| Male sex | 5068 (70.53%) | 1.01 | 0.89-1.14 | 0.93 | 0.91 | 0.79-1.05 | 0.2 |
| Age (years) | 64 (57-71) | 2.52 | 2.23-2.84 | <10<sup>-50</sup> | 2.63 | 2.27-3.04 | <10<sup>-37</sup> |
| BMI (kg/m<sup>2</sup>) | 28.3 (25.8-31) | 0.91 | 0.8-1.03 | 0.15 | 0.85 | 0.73-1 | 0.05 |
| History of MI | 1549 (21.56%) | 1.16 | 1.02-1.31 | 0.02 | 1.23 | 1.06-1.42 | <10<sup>-2</sup> |
| CABG | 199 (2.77%) | 1 | 0.73-1.37 | 0.99 | 1.04 | 0.72-1.51 | 0.83 |
| Previous PCI | 1883 (26.20%) | 0.91 | 0.81-1.03 | 0.15 | 0.87 | 0.75-1.01 | 0.07 |
| Stroke/TIA | 490 (6.82%) | 2.01 | 1.69-2.38 | <10<sup>-14</sup> | 2.04 | 1.66-2.51 | <10<sup>-10</sup> |
| Atrial fibrillation | 820 (11.41%) | 2.07 | 1.79-2.38 | <10<sup>-22</sup> | 2.25 | 1.91-2.66 | <10<sup>-20</sup> |
| PAD | 780 (10.85%) | 2 | 1.74-2.3 | <10<sup>-21</sup> | 1.91 | 1.61-2.26 | <10<sup>-12</sup> |
| CKD | 509 (7.08%) | 3.52 | 3.03-4.1 | <10<sup>-59</sup> | 3.87 | 3.25-4.62 | <10<sup>-50</sup> |
| VHD | 438 (6.10%) | 2.19 | 1.84-2.6 | <10<sup>-18</sup> | 2.33 | 1.9-2.85 | <10<sup>-15</sup> |
| DCM | 600 (8.35%) | 1.95 | 1.66-2.29 | <10<sup>-15</sup> | 2.22 | 1.85-2.67 | <10<sup>-16</sup> |
| COPD | 633 (8.81%) | 2.32 | 2.01-2.69 | <10<sup>-28</sup> | 2.43 | 2.04-2.89 | <10<sup>-22</sup> |
| Diabetes Mellitus | 1441 (20.05%) | 1.49 | 1.32-1.68 | <10<sup>-10</sup> | 1.44 | 1.25-1.67 | <10<sup>-5</sup> |
| HBP | 3788 (52.71%) | 1.17 | 1.03-1.31 | 0.01 | 1.15 | 1-1.33 | 0.05 |
| Dyslipidaemia | 2973 (41.37%) | 0.98 | 0.87-1.09 | 0.7 | 0.91 | 0.8-1.04 | 0.19 |
| LVEF (%) | 50 (40-55) | 0.46 | 0.4-0.52 | <10<sup>-27</sup> | 0.39 | 0.33-0.46 | <10<sup>-25</sup> |
| NTproBNP(pg/ml) | 932(206-2635) | 3.33 | 1.93-5.76 | <10<sup>-4</sup> | 2.2 | 1.23-3.94 | <10<sup>-2</sup> |
| Creatinine(mg/dL) | 0.92 (0.8-1.11) | 1.86 | 1.66-2.09 | <10<sup>-25</sup> | 2.02 | 1.76-2.33 | <10<sup>-22</sup> |
| HGB (g/dL) | 13.8 (12.6-14) | 0.55 | 0.49-0.62 | <10<sup>-21</sup> | 0.54 | 0.47-0.62 | <10<sup>-16</sup> |
| PLT (×10<sup>3</sup>/μL) | 226 (188-271) | 1.01 | 0.9-1.13 | 0.84 | 0.97 | 0.85-1.11 | 0.68 |
| WBC (×10<sup>3</sup>/μL) | 8.1 (6.6-10.1) | 1.27 | 1.13-1.42 | <10<sup>-4</sup> | 1.32 | 1.15-1.51 | <10<sup>-4</sup> |
| STEMI | 1721 (23.95%) | 1.29 | 1.13-1.46 | <10<sup>-3</sup> | 1.38 | 1.19-1.61 | <10<sup>-4</sup> |
| NSTE-MI | 593 (8.25%) | 1.73 | 1.46-2.07 | <10<sup>-9</sup> | 1.91 | 1.56-2.34 | <10<sup>-9</sup> |
| UA | 1377 (19.16%) | 0.96 | 0.83-1.1 | 0.52 | 0.9 | 0.76-1.07 | 0.22 |
| CCS | 3495 (48.64%) | 0.72 | 0.64-0.8 | <10<sup>-8</sup> | 0.68 | 0.59-0.77 | <10<sup>-7</sup> |

BMI – body mass index; CABG – coronary artery bypass graft; CCS – chronic coronary syndrome; CI – confidence interval; CKD – chronic kidney disease; COPD – chronic obstructive pulmonary disease; CV – cardiovascular; DCM – dilated cardiomyopathy; HGB – haemoglobin; HBP – high blood pressure; LVEF – left ventricular ejection fraction; MI – myocardial infarction; PAD – peripheral artery disease; PCI – percutaneous coronary intervention; PLT – platelet count; NSTE-MI – non-ST segment elevation acute MI; STEMI – ST segment elevation acute MI; TIA – transient ischemic attack; UA – unstable angina; VHD – valvular heart disease; WBC – white blood cells count.

### Machine learning analysis

Machine learning models accurately predicted out-of-hospital survival at all timeframes on all time-dependent performance metrics (Figure 3). While AUC-ROC was consistently high (Figure 3A, 3C), AUC-PR gradually increased as the deceased-to-alive ratio became less imbalanced (Figure 3B, 3D). Minor differences were between testing dataset predictions and validations dataset predictions, revealing that overfitting did not occur (Figure 3). Tuned hyperparameters found by Bayesian search included a total of 5000 aggregated decision trees, with a maximum tree depth of 12 levels and a learning rate of 0.01 (see supporting code). Most important survival predictors were obtained by classifying the averaged importance position at all timeframes. For all-cause mortality, left ventricular ejection fraction (LVEF), age, hospitalization cost, heart rate at presentation, renal function reflected by maximum creatinine during hospitalization, standard deviation of red cell distribution width and last determined before discharge lymphocyte per monocyte ratio were among the most important parameters. For cardiovascular cause mortality, left ventricular ejection fraction (LVEF), age, systolic and diastolic blood pressure at presentation, heart rate at presentation, hospitalization cost, renal function reflected by maximum creatinine during hospitalization, the standard deviation of red cell distribution width and last determined before discharge lymphocyte per monocyte ratio was among the most important parameters. Furthermore, the most important factors for the occurrence of adverse events and most important factors against the occurrence of adverse events of any cause and cardiovascular cause are reported in Supplemental Figure 1. Time-dependent SHAP values are reported in Figure 5 and reveal how the predictor’s values (e.g., advanced age or reduced LVEF) predispose to adverse events. Patient individualized survival prediction can be illustrated using ML probability output at each timeframe with event occurrence estimated at the point where probability is less than the cut-off for that specific timeframe (Figure 6).

**Figure 3:**
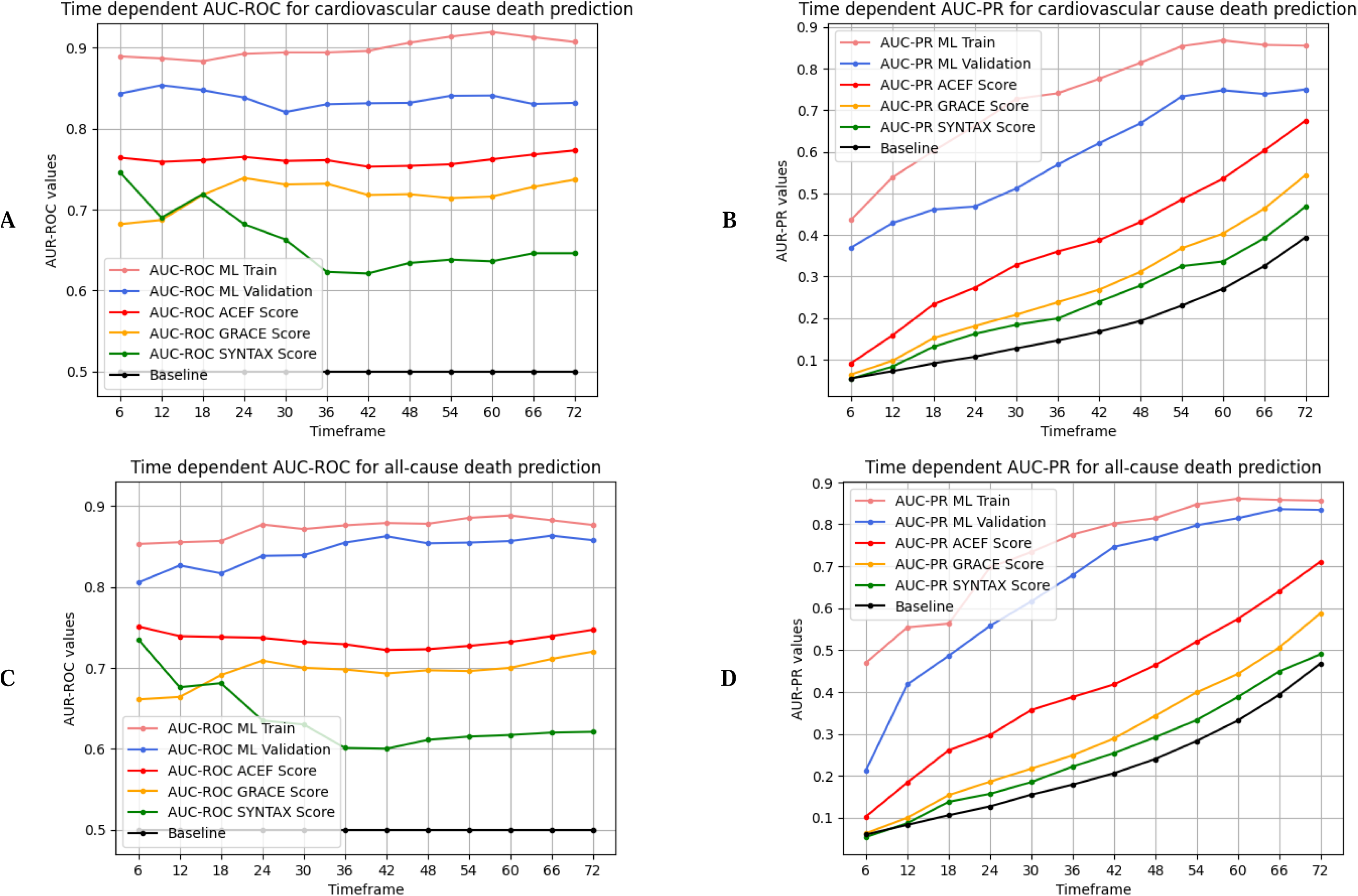
Time-dependent AUC-ROC and AUC-PR for cardiovascular cause and all-cause death prediction.

### Comparison between machine learning and clinical scores

To illustrate the predictive performance of ML models, some important clinical risk scores, such as ACEF, GRACE, or SYNTAX scores, were also calculated and compared with ML models. ML consistently and significantly outperformed clinical risk scores on all timeframes (Figure 3) and all performance metrics (Table 3). Among clinical scores, the ACEF score had the highest performance metrics, while the SYNTAX score had the lowest performance metrics. The integrated AUC-ROC for prediction of all-cause mortality by the ML model versus ACEF score was 0.844 and 0.735, respectively (Table 3). The integrated AUC-ROC for prediction of cardiovascular cause mortality by the ML model versus ACEF score was 0.837 and 0.761, respectively (Table 3). A more striking increase in prediction performance is reflected by the integrated AUC-PR and integrated baseline-corrected AUC-PR. The integrated AUC-PR for prediction of cardiovascular and all-cause mortality by the ML model was 0.647 and 0.589, respectively, while for ACEF score was 0.380 and 0.410, respectively (Table 3). The integrated baseline corrected AUC-PR for prediction of cardiovascular and all-cause mortality by the ML model was 0.407 and 0.428, respectively, while for ACEF score was 0.199 and 0.190, respectively (Table 3). Additional details regarding performance metrics are reported in Supplemental Table 3. Other metrics, such as integrated MCC, F1, and Brier score were also in favor of ML models.

**Table 3:** Performance metrics for ML models and clinical risk scores.

| Parameter | Integrated AUC-ROC* |  | Integrated AUC-PR* |  | Integrated cAUC-PR | Integrated MCC | Integrated F1 score | Integrated Brier score |
| --- | --- | --- | --- | --- | --- | --- | --- | --- |
| Cardiovascular cause death prediction |  |  |  |  |  |  |  |  |
| ML on training dataset | 0.900 | - | 0.727 | - | 0.546 | 0.583 | 0.664 | 0.094 |
| ML on validation dataset | 0.837 | - | 0.589 | - | 0.407 | 0.438 | 0.555 | 0.117 |
| ACEF score | 0.761 | p<10<sup>-10</sup> | 0.380 | p<10<sup>-10</sup> | 0.199 | 0.282 | 0.351 | 0.124 |
| GRACE score | 0.718 | p<10<sup>-10</sup> | 0.275 | p<10<sup>-10</sup> | 0.093 | 0.231 | 0.301 | 0.159 |
| SYNTAX score | 0.662 | p<10<sup>-10</sup> | 0.238 | p<10<sup>-10</sup> | 0.238 | 0.141 | 0.216 | 0.173 |
| All-cause death prediction |  |  |  |  |  |  |  |  |
| ML on training dataset | 0.873 | - | 0.736 | - | 0.517 | 0.543 | 0.650 | 0.107 |
| ML on validation dataset | 0.844 | - | 0.647 | - | 0.428 | 0.485 | 0.612 | 0.125 |
| ACEF score | 0.735 | p<10<sup>-10</sup> | 0.410 | p<10<sup>-10</sup> | 0.190 | 0.278 | 0.387 | 0.150 |
| GRACE score | 0.695 | p<10<sup>-10</sup> | 0.295 | p<10<sup>-10</sup> | 0.075 | 0.220 | 0.328 | 0.175 |
| SYNTAX score | 0.637 | p<10<sup>-10</sup> | 0.255 | p<10<sup>-10</sup> | 0.036 | 0.137 | 0.241 | 0.186 |

ACEF – Age, Creatinine and Ejection Fraction score; AUC-ROC – area under the curve of receiver-operator characteristic; AUC-PR – area under the curve of precision-recall; cAUC-PR – baseline corrected area under the curve of precision-recall; GRACE – Global Registry of Acute Coronary Events; MCC – Matthews correlation coefficient; ML – machine learning; SYNTAX – Synergy Between PCI with TAXUS and Cardiac Surgery. \*p values were obtained by comparing each clinical score with ML on validation dataset at each timeframe.

## Discussions

The main findings of our study are: (1) In a prospective all-comers PCI registry, both all-cause and cardiovascular cause out-of-hospital mortality is high, with 6-years mortality of up to 42%; (2) ML models can accurately predict long-term out-of-hospital survival in a CAD population treated by PCI using routine laboratory parameters; (3) Predictive performance of ML models was better than relevant clinical scores, such as GRACE, ACEF or SYNTAX score on all performance metrics, including integrated AUC-ROC, AUC-PR, MCC, F1 and Brier score; (3) Important factors for survival prediction includes age, LVEF, blood pressure and heart rate on presentation. However, most important factors were those derived from laboratory parameter analysis; (4) Time-dependent SHAP values reveal how the values of a parameter impact predictions for each timeframe.

Ischemic heart disease is the leading cause of death^14^, although precise long-term follow-up from the moment of CAD diagnosis, especially out-of-hospital, is scarce. In our study 5-year and 6-year all-cause, out-of-hospital mortality was 21.7% and 33.2%, respectively, relatively lower than the 5-year all-cause mortality of 37.3% from the SYNTAX trial although it included only three-vessel CAD disease.^15^ In patients with established CAD, clinical risk prediction tools for secondary prevention are useful, considering the high mortality associated with CAD. For this purpose, certain clinical scores such as ACEF, SYNTAX, and GRACE scores were developed.^1,2,4^ Refinements of clinical scores were attempted to improve predictive performance, particularly for the original SYNTAX score, from which several SYNTAX-derived scores were reported.^16^ In our analysis, ML models outperformed clinical scores on all timeframes and all statistical metrics (Table 3). Accordingly, the use of machine learning instead of continuously refined traditional risk scores seems to be more efficient in long-term mortality risk prediction in CAD patients, especially if clinical data can reliantly be obtained from EHR. In the present study, over 600,000 laboratory values were automatically obtained from the EHR, making future integration in clinical practice possible.

In our analysis, LVEF, age, blood pressure, and heart rate were among the most important predictors, which are also partially included in clinical scores such as ACEF and GRACE scores. High hospitalization cost and increased contrast volume were also predictors of events, reflecting the need for advanced therapies and long or complex PCI procedures, respectively. Interestingly, numerous important clinical features were obtained from laboratory values, reflecting renal function (first and maximum serum creatinine and urea levels), hematologic function (red cell distribution width, mean corpuscular hemoglobin concentration, platelet distribution width), inflammatory status (lymphocyte per monocyte ratio, maximum neutrophil count) and glycemic status (Figure 4). Most of the mentioned parameters were previously reported as predictors of adverse events. The association between lymphocyte-to-monocyte ratio and impaired prognosis in the CAD population has been documented in a recent meta-analysis.^17^ Other studies showed that red cell distribution width is an inexpensive prognostic marker in both myocardial infarction and heart failure.^18,19^ This illustrates that precision phenotyping can be achieved using laboratory parameters. Moreover, while the clinician works with dichotomized outcomes from continuous variables (e.g., patient has heart failure with reduced ejection fraction if LVEF ≤40%) leading to an information loss^20^, ML models are particularly powerful in analyzing continuous data without the need for fitting into categories – for this reason, the most important parameters were laboratory values instead of disease labels. Interestingly, the severity of coronary lesions was not particularly important for long-term prognosis. Similarly, SYNTAX score performance decreased over time, revealing that coronary anatomy is less important in the long term.

**Figure 4:**
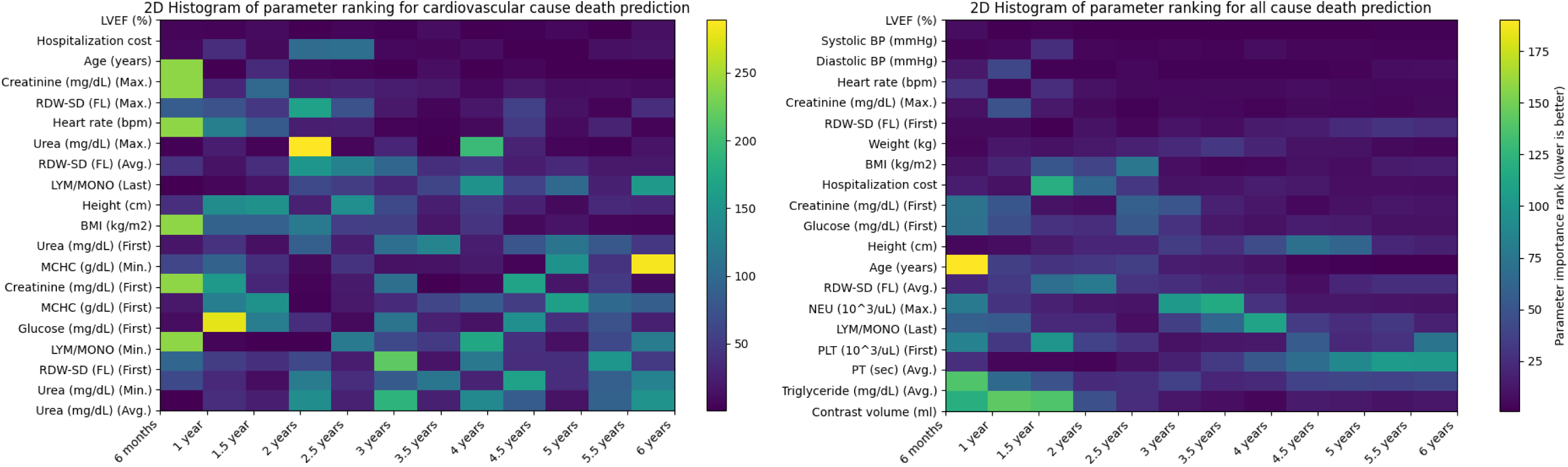
Most important parameters in the prediction of cardiovascular cause and all-cause mortality (lower is better).

**Figure 5:**
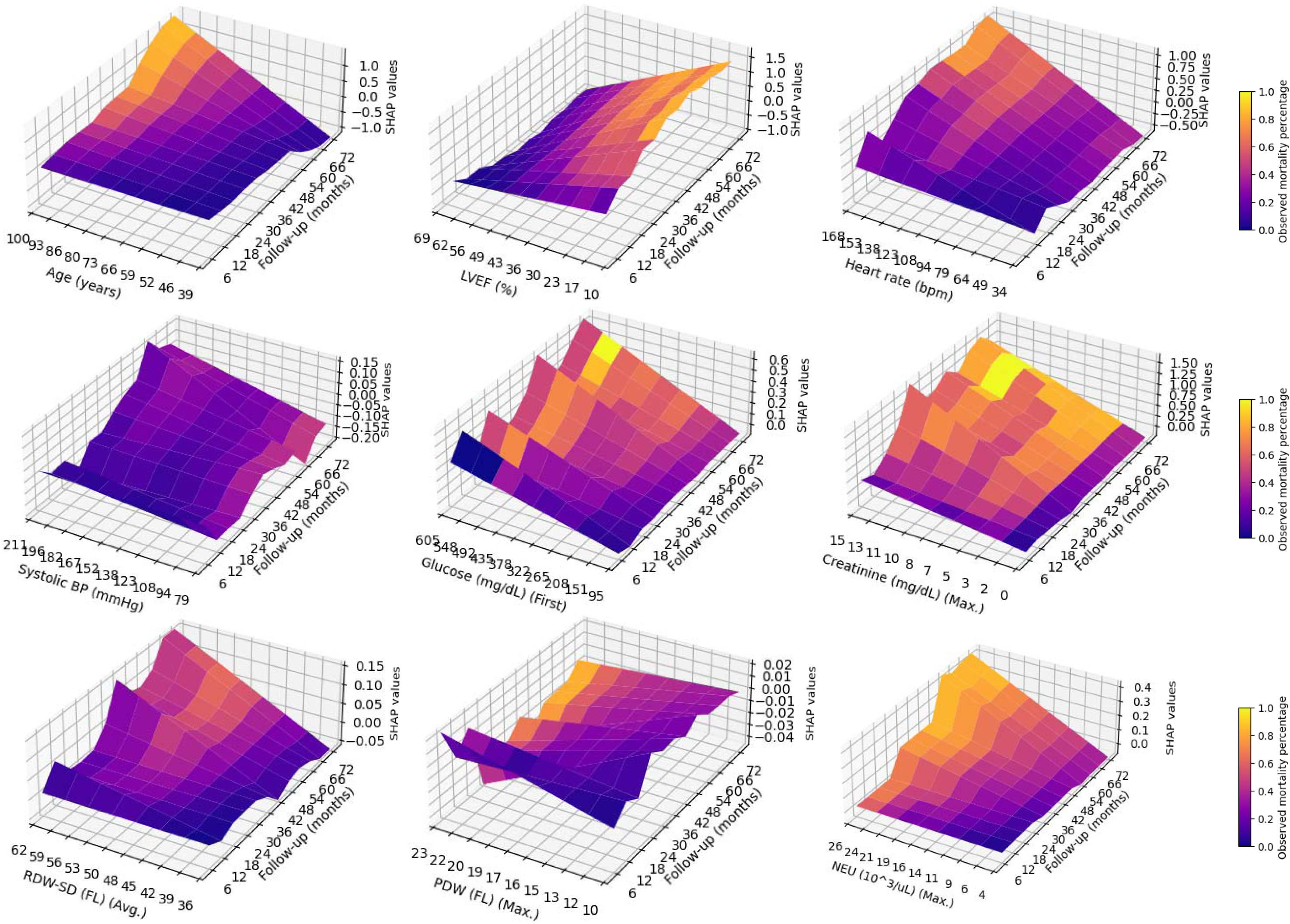
4D time-dependent SHAP mesh plot for various predictors of all-cause mortality.

**Figure 6:**
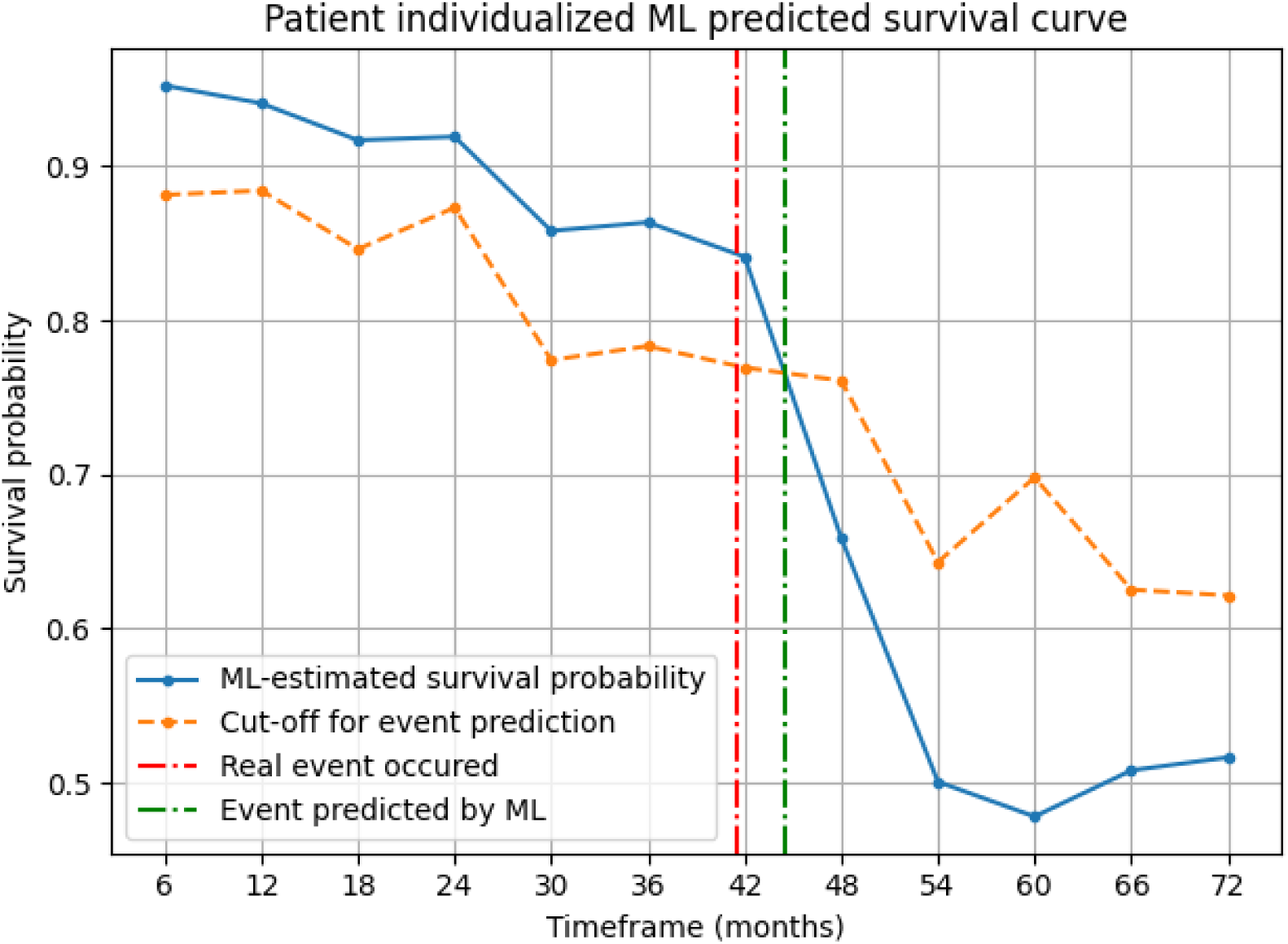
An example of a patient specific estimated survival curve.

The emergence of AI brought numerous tools for prediction making, which can be simplistically categorized as supervised learning using machine learning techniques (e.g., support vector machines or gradient-boosted decision trees) or unsupervised learning using deep learning techniques (e.g., neural networks). Depending on the underlying task, certain algorithms could be more suitable. Indeed, for tabular data, deep learning seldom outperforms machine learning.^21^ Regarding survival analysis, Kaplan–Meier and Cox proportional hazards models have successfully been used for decades.^22^ However, both techniques consider a linear relationship between the risk factor and the log hazard function^23^. This constant effect of the risk factor during the follow-up assumed by the Kaplan–Meier and Cox models could be an oversimplification.^23^ Recently, ML methods were proposed to analyze survival data.^24^ More specifically, standard ML techniques use binary classification to predict the outcome at a certain timeframe. This requires dichotomization with removal of censored data, does not consider hazards a function of time, but has a high interpretability of the model. On the other hand, modified ML techniques for survival data are adjusted to consider hazards as a function of time, to handle censored and time-to-event data, but have lower interpretability.^24^ Moreover, numerous performance metrics to evaluate survival prediction models are time-dependent and require the removal of censored events (e.g., Brier score or time-dependent area under receiver operator characteristic).^25^ Indeed, there are a variety of ML and deep learning techniques to analyze survival data, and the best technique to perform survival analysis is yet to be determined.

A reproducibility crisis is increasingly recognized in the area of AI, driven mainly by data leakage and lack of transparency by unpublished code.^26,27^ Indeed, data leakage – the ability of the AI model to see the outcome of the training dataset – is an important problem that may remain undetected.^28^ In our previous study, an unexpected data leak was observed – the ML model was making an unrealistic nearly perfect in-hospital mortality prediction by looking at treatment at discharge, recognizing that if a patient did not have medical treatment at hospital discharge it was because the patient was deceased.^29^ In the current study, we consider data leak to be highly improbable, since (1) only in-hospital data was analyzed, (2) only out-of-hospital death was considered, and (3) survival data was acquired from a different institution. Moreover, survival data was not merged with clinical data during the analysis. Regarding underlying code, a report revealed that 6% of AI papers are accompanied by complete code, while half of the AI papers contained “pseudocode”, a brief description of the algorithm.^30^ In line with current recommendations, the underlying algorithm code for the current study is publicly disclosed.

### Study limitations and future research directions

The main limitation of our study is the lack of external validation. In addition, the study population is typical for Eastern Europe, thus extrapolating to other populations could be limited. Although not a limitation per se, we are the first to illustrate a time-dependent SHAP plot and certain time-integrated statistical indicators (e.g., integrated AUC-PR and baseline corrected AUC-PR) which warrants further investigation.

## Conclusions

Machine learning can accurately predict out-of-hospital all-cause and cardiovascular cause death using routinely performed laboratory parameters and outperforms classic clinical parameter-based risk scores in a prospectively followed CAD population treated by PCI.

## Acknowledgments

The authors would like to thank all the resident colleagues who continuously introduced the data regarding PCIs performed in the Emergency Institute for Cardiovascular Diseases and Transplantation since 2016. We also would like to thank Mr. Ioan Matei for his help in achieving long-term mortality data.

## Author contributions

P.-A.C. and L.H. were involved in the study design. P.-A.C. and L.H. performed the literature search. L.H. supervised and coordinated the data collection in the registry. P.-A.C. was involved in the development of machine learning models and explainable artificial intelligence. All authors were involved in data collection. T.O. was responsible for the development and maintenance of the online platform. P.-A.C. and L.H. analyzed the data. P.-A.C. and L.H. were involved in the writing of the initial draft. All authors were involved in the writing of the final draft and approved the manuscript.

## Data availability

Due to national and EU regulations, particularly the General Data Protection Regulation, the data used in this study cannot be made publicly available and shared with the wider research community. The data can be shared by the corresponding author for use in secure environments upon request. However, all Python scripts underlying this study are publicly available at https://github.com/paul-md/Survival-ML.

